# The Heartbeat Study: Feasibility and Advertisement Costs of Implementing a Digital Strategy to Enhance Diversity in the LIBREXIA-AF Clinical Trial

**DOI:** 10.64898/2026.08.24.26361277

**Authors:** Tasmeen Hussain, Yixin Wang, Ying Chen, Garrick Olson, Boris Panitch, Kara Clemins, Nadia Elkarra, Karma Lhamo, Nicole Odenwald, Sneha Jain, David Hufner, Madeline Quall, Christine Anderson, Marco Perez

**Affiliations:** Division of Cardiovascular Medicine (Clinical Cardiac Electrophysiology), Department of Medicine, Stanford University, Stanford, CA; Stanford Prevention Research Center, Stanford University, Stanford, CA; Technology and Digital Solutions, Stanford University, Stanford, CA; Division of Hospital Medicine, Stanford University, Stanford, CA; Stanford Center for Clinical Research, Stanford University, Stanford, CA; Division of Cardiovascular Medicine, Department of Medicine, Stanford University, Stanford, CA; Johnson & Johnson Innovative Medicine, New Brunswick, NJ

## Abstract

**Background:** Recruitment of diverse participants remains a challenge in cardiovascular clinical trials. Little is known about how recruitment efficiency and advertising costs with web-based tools vary across US communities. We evaluated an online recruitment platform and examined the cost of acquiring both all-comers and diverse participants in relation to community-level income.

**Methods:** The Heartbeat Study evaluated a digital recruitment strategy to identify US participants for the ongoing Phase 3 LIBREXIA-AF trial. Online advertisements directed individuals with atrial fibrillation to a pre-screening website, where demographic and health data were collected. Advertising impressions, clicks, and costs were recorded. Participant ZIP codes were linked to Core Based Statistical Areas (CBSAs) and CBSA-level income. We measured recruits from underrepresented groups (women, African Americans, Latinos) completing online registration per $100,000 in advertising expenses. Click-weighted linear regression evaluated associations between CBSA income and advertising efficiency.

**Results:** A total of 1,406 recruits completed online registration, with 1,319 participants from 260 CBSAs included in the geographic analysis. Participants were 73 years old on average; 547 (41.5%) were women, 59 (4.5%) African American, and 44 (3.3%) Latino. A total of $163,949.13 was spent on 82,681,711 impressions and 454,750 clicks. Recruits per $100,000 in advertising spend were 334 for women, 36 for African Americans, and 27 for Latinos. CBSA-level income was modestly inversely associated with cost per impression (R^2^=0.058; p<0.001) and cost per click (R^2^=0.038; p=0.005), but not recruitment yield for African Americans (p=0.99), Latinos (p=0.37), or women (p=0.21) (R^2^ range, 0.000– 0.13).

**Conclusion:** In this national analysis, online advertising enabled broad engagement across diverse US communities, but income was not associated with recruitment yield among women, African American, or Latino participants. Minority representation remained limited, suggesting digital recruitment alone may be insufficient to improve trial diversity. Targeted, culturally and linguistically tailored strategies may be needed to enhance diverse recruitment.

## Introducion

The recruitment of diverse patients remains one of the most significant challenges in modern clinical trial conduct.^1^ Participants from certain populations, including various racial and ethnic groups and women, continue to be largely underrepresented in cardiovascular clinical trials.^2–6^ These disparities could potentially limit the generalizability of trial findings and constrain the development of broadly applicable evidence-based therapies.

Digital recruitment strategies, including online, mobile, and virtual approaches, may directly address several challenges faced by underrepresented populations.^7–9^ Online recruitment helps to limit travel costs, allows participation of those with mobility challenges, and extends reach to rural patient populations. Prior studies have shown that digital advertising can effectively support clinical trial recruitment; information and communication platforms have improved the intent of diverse participants to enroll in clinical trials,^10^ facilitated crowdsourcing for enhanced trial design,^11,12^ and have enabled exceptional patient reach.^13,14^ On the other hand, digital recruitment may select for younger, more educated, and more technologically savvy participants,^15^ suggesting a digital divide^16^ which requires continued attention.

There remains limited guidance for the optimization of diversity in clinical trials. The US Food and Drug Administration (FDA) currently has nonbinding recommendations for Phase 3 trials which support the development of diversity action plans which outline the sponsor’s inclusion goals for age, sex, race, and ethnicity of their study population as well as rationale and plans to reach those goals.^17^ As non-White populations are expected to become the majority in the US by 2045,^18^ building diverse representation into current trials is critical. US communities are heterogeneous in terms of income, race, ethnicity, education and other factors; it is important to consider how these regional socioeconomic characteristics may influence advertising costs, participant engagement, and trial enrollment patterns.

Research teams continue to explore how digital recruitment may be optimized to help reach equity goals in research. However, the majority of studies in this space focus on the effectiveness of specific digital marketing strategies such as social media^19^ and search engine optimization.^20^ No large analysis has specifically examined how digital recruitment varies across US communities of different demographic and income breakdowns; city-centric data would help improve advertising quality and diverse recruitment in cardiovascular clinical trials.

To better understand the feasibility and advertisement costs to recruit diverse US participants, we performed the Heartbeat Study, which utilized a digital advertising platform to recruit a subset of patients to a cardiovascular clinical trial (LIBREXIA-AF). We also described how advertising to specific community populations of differing income-levels affected total and diverse (African American, Latino, and women) participant recruitment.

## Methods

### Digital Platform

The Heartbeat Study was an observational study designed to evaluate a digital platform used for recruitment for US participants to the LIBREXIA-AF clinical trial. LIBREXIA-AF is an ongoing, international phase 3 study designed to evaluate the safety and efficacy of a novel factor XIa inhibitor, milvexian, for stroke and systemic embolism prevention in atrial fibrillation. The full protocol for LIBREXIA-AF and its trial registration data have been previously published.^21,22^ The Heartbeat Study was approved by the Stanford University Institutional Review Board. All participants provided informed consent prior to completing the web-based registration.

The Stanford Center for Clinical Research served as the clinical and data coordinating center and oversaw development of the web-based platform used for participant pre-screening and data capture. A contract research organization (IQVIA Holdings, Inc; Durham, NC), supported coordination with clinical trial sites. A marketing agency (CT Media; New York, NY) developed targeted digital advertising campaigns focused on geographic targets to direct individuals with atrial fibrillation to the Heartbeat Study pre-screening platform. Advertising campaigns and marketing strategy were funded by Bristol Myers Squibb and Johnson & Johnson with additional recommendations provided by the research and technical teams at Stanford Health Care.

Advertisements were delivered via online social media (ex. Facebook), search engines including Google, Bing and Yahoo, as well as native websites. Ads were accessible by mobile phone, desktop, and tablet. Selected images of our advertisements can be found in Supplement 1. Due to limited ability to directly target audiences based on demographics across media platforms, the campaign utilized female-focused imagery as the primary method to engage female audiences. In addition, most of the patients displayed in the advertisements were African American. Search engine terms and frequency of searched terms can be found in Supplement 2. The most searched terms we targeted through our campaign were “afib,” “atrial fibrillation,” and “af arrhythmia,” each with over 160,000 average monthly searches. Social media and banner ad copy (text) is shown in Supplement 3. Ads were varied and focused on addressing personal health risks (“Atrial fibrillation can lead to blood clots, heart attacks and strokes”) as well as the need for geographic and racial/ethnic diversity in atrial fibrillation trials (“People from diverse backgrounds can help make a difference together by participating in Afib research.”)

Eligible individuals who clicked on online advertisements were directed to a web-based application to complete a pre-screening questionnaire. Participant-collected variables included demographics such as age, sex at birth, and self-reported ethnicity as well as the presence of one or more of sixteen comorbid health conditions. The full survey (including screenshots) and website flow diagram can be found in Supplement 4. Those who completed the survey were evaluated for LIBREXIA-AF trial eligibility and were offered referrals to participating LIBREXIA-AF clinical trial sites for further screening before LIBREXIA-AF trial participation. Participants were eligible for the Heartbeat Study analysis if they were at least 18 years of age, reported a diagnosis of atrial fibrillation, and were able to complete the Heartbeat Study web-based survey in English. Eligibility for participation in the LIBREXIA-AF clinical trial was assessed separately according to the trial’s prespecified inclusion and exclusion criteria.^23^

### Data Collection

Participant data were collected through a web application developed by Stanford Research Technology and Digital Solutions and stored within the Stanford CHOIR system, a HIPAA-compliant platform designed for secure storage of protected health information. Advertising metrics, including impressions (the number of times an advertisement was displayed), clicks, click-through-rate (CTR; clicks/impression x 100) and total advertising costs were collected by CT Media. Geographic data were used to link participant zip codes to U.S. Core-Based Statistical Areas (CBSAs).^24–26^ We obtained CBSA-level income data using publicly available U.S. Census and Bureau of Economic Analysis datasets.^27^ We developed a study-specific metric (“diverse recruitment yield”) to estimate each area’s potential to recruit diverse participants (e.g., women, African Americans, Latinos) by calculating the number of diverse participants recruited per $100,000 of advertising expenditure. This was calculated by $100,000/total advertising cost to a CBSA in dollars * number of diverse recruits. To limit bias, we only included CBSAs in this analysis with more than 20 total recruits.

### Statistical Analysis

The primary endpoint was the estimated number of participants recruited to the Heartbeat Study per $100,000 spent on advertisement from different CBSAs. Secondary endpoints included the number of participants who completed the Heartbeat Study web-based registration, the number of participants identified as eligible for enrollment in the LIBREXIA-AF clinical trial and the number of participants referred to LIBREXIA-AF from prespecified diversity categories, including women, African Americans, and Latinos across CBSAs. Additional endpoints included measures of advertising resources utilized, including cost per click and impressions per click across CBSAs.

Continuous variables were presented as mean ± standard deviation or median with interquartile range as appropriate, and categorical variables were presented as counts and percentages. Linear regression models were used to evaluate the relationship between CBSA-level income and advertising resource utilization, including cost per click, impressions per click and recruits/$100,000 spent. Statistical analyses were performed using R (R Foundation for Statistical Computing, Vienna, Austria). A two-sided significance level of 0.05 was used.

## Results

### Participant Recruitment

Recruitment to the Heartbeat Study was performed from December 18, 2024 to February 11, 2025. A total of 1,406 participants completed the recruitment registration. A total of $163,949.13 were spent for a total of 82,102,081 impressions, 452,691 clicks and 0.55% CTR. Demographic and health information of all participants who completed the registration survey are summarized in Table 1. The average cost per recruit was $116.61. The average age of participants was 73 years old; 575 (41%) were women; 1,201 (85%) were white, 62 (4%) were African American, 35 (2%) were Latino, 38 (3%) were Asian and 47 (3%) noted other or mixed race. Participants who qualified for the LIBREXIA-AF clinical trial were more likely to be older than non-qualifying patients (p < 0.001). Those who qualified were more likely to have hypertension and atherosclerosis than non-qualifiers (p <0.05) but equally likely to have had strokes or diabetes. Significantly more non-qualifiers than qualifiers had blood clots, liver disease, heart valve replacements or were on dialysis (p > 0.05).

**Table 1:**
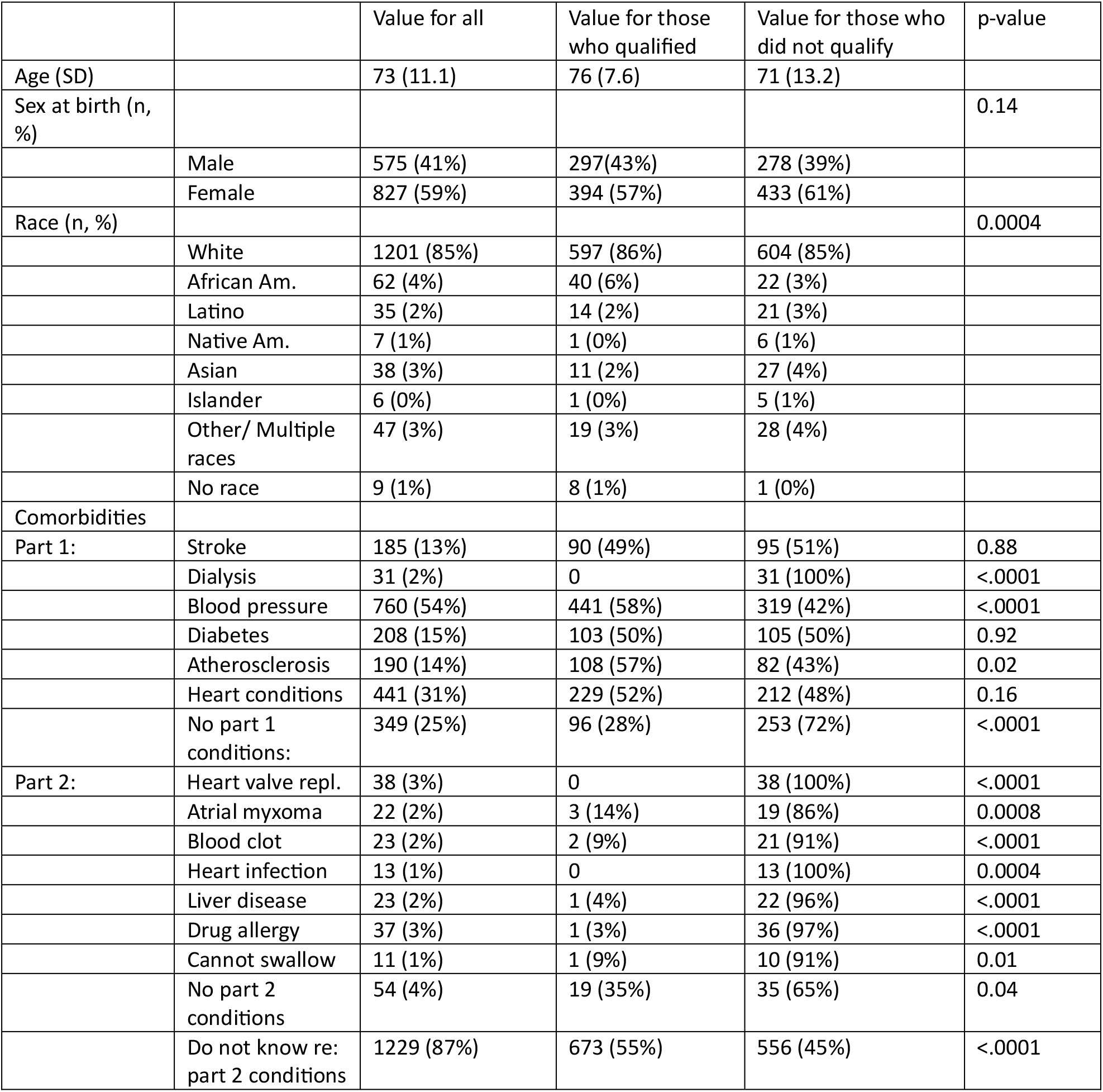
Baseline characterisics by qualificaion for the LIBREXIA-AF study.

|  |  | Value for all | Value for those who qualified | Value for those who did not qualify | p-value |
| --- | --- | --- | --- | --- | --- |
| Age (SD) |  | 73 (11.1) | 76 (7.6) | 71 (13.2) |  |
| Sex at birth (n, %) |  |  |  |  | 0.14 |
|  | Male | 575 (41%) | 297(43%) | 278 (39%) |  |
|  | Female | 827 (59%) | 394 (57%) | 433 (61%) |  |
| Race (n, %) |  |  |  |  | 0.0004 |
|  | White | 1201 (85%) | 597 (86%) | 604 (85%) |  |
|  | African Am. | 62 (4%) | 40 (6%) | 22 (3%) |  |
|  | Latino | 35 (2%) | 14 (2%) | 21 (3%) |  |
|  | Native Am. | 7 (1%) | 1 (0%) | 6 (1%) |  |
|  | Asian | 38 (3%) | 11 (2%) | 27 (4%) |  |
|  | Islander | 6 (0%) | 1 (0%) | 5 (1%) |  |
|  | Other/ Multiple races | 47 (3%) | 19 (3%) | 28 (4%) |  |
|  | No race | 9 (1%) | 8 (1%) | 1 (0%) |  |
| Comorbidities |  |  |  |  |  |
| Part 1: | Stroke | 185 (13%) | 90 (49%) | 95 (51%) | 0.88 |
|  | Dialysis | 31 (2%) | 0 | 31 (100%) | <.0001 |
|  | Blood pressure | 760 (54%) | 441 (58%) | 319 (42%) | <.0001 |
|  | Diabetes | 208 (15%) | 103 (50%) | 105 (50%) | 0.92 |
|  | Atherosclerosis | 190 (14%) | 108 (57%) | 82 (43%) | 0.02 |
|  | Heart conditions | 441 (31%) | 229 (52%) | 212 (48%) | 0.16 |
|  | No part 1 conditions: | 349 (25%) | 96 (28%) | 253 (72%) | <.0001 |
| Part 2: | Heart valve repl. | 38 (3%) | 0 | 38 (100%) | <.0001 |
|  | Atrial myxoma | 22 (2%) | 3 (14%) | 19 (86%) | 0.0008 |
|  | Blood clot | 23 (2%) | 2 (9%) | 21 (91%) | <.0001 |
|  | Heart infection | 13 (1%) | 0 | 13 (100%) | 0.0004 |
|  | Liver disease | 23 (2%) | 1 (4%) | 22 (96%) | <.0001 |
|  | Drug allergy | 37 (3%) | 1 (3%) | 36 (97%) | <.0001 |
|  | Cannot swallow | 11 (1%) | 1 (9%) | 10 (91%) | 0.01 |
|  | No part 2 conditions | 54 (4%) | 19 (35%) | 35 (65%) | 0.04 |
|  | Do not know re: part 2 conditions | 1229 (87%) | 673 (55%) | 556 (45%) | <.0001 |

After removing those participants without complete personal and/or CBSA data, a total of 1319 remained in the analysis from 260 unique CBSAs. Of those patients, the average age was 73 years old, 547 (41.5%) were women, 59 (4.5%) were African American and 44 (3.3%) were Latino (or of mixed race including either ethnic group). The CBSAs with the most survey representation included New York-Newark-Jersey City, NY-NJ (n = 113), Los Angeles-Long Beach-Anaheim, CA (n = 95), and Washington-Arlington-Alexandria, DC-VA-MD-WV (n = 66). Of the 691 participants pre-qualified for LIBREXIA-AF, 11 declined referral to a study site for further screening for LIBREXIA-AF.

### Advertisement Costs

A total of $163,949.13 were spent on online advertisement for the Heartbeat Study for a total of 82,681,711 impressions and 454,750 clicks (CTR 0.55). Over 1,406 recruits, this represented a cost/participant of $116.61. This cost was split amongst advertisements on search engines, which received $47,754 for 395,405 impressions and 52,582 clicks (CTR 13), social media advertising, which received $67,476 for 8,674,048 impressions and 134,647 clicks (CTR 1.6) and native websites, which received $48,719 for 73,612,258 impressions and 267,521 clicks (CTR 0.36). In terms of advertising yield, in total of $163,949.13 was spent to recruit 1406 unique participants, with a recruits/$100K advertising spend of 857 for all-comers. The number of estimated recruits/$100K was 334 for women, 36 for African Americans and 27 for Latinos.

### Advertisement Yield by CBSA

Higher CBSA-level income was associated with lower advertising cost per impression (R^2^ = 0.058; p < 0.001) and cost per click (R^2^ = 0.038; p = 0.005) in linear regression analyses weighted by clicks. (Figures 1a and 1b). Two parallel tiers of communities with Cost per Impression by CBSA Income were evident (Figure 1a) As we weighted by clicks, the regression line closely mirrored the lower tier.

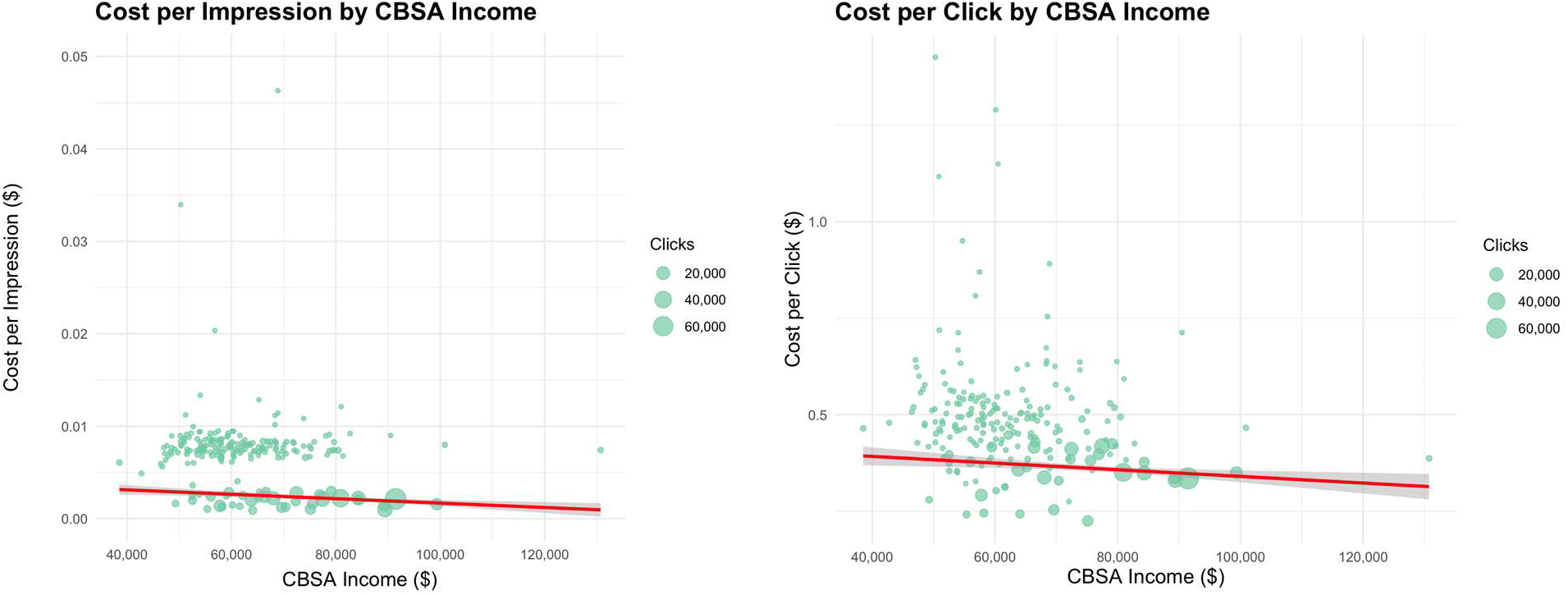
Figure 1a: Cost per Impression by CBSA Income and Figure 1b: Cost per Click by CBSA (red line represents linear regression fit)

Finally, we evaluated whether CBSA-level income was associated with the estimated diverse recruits/$100K advertising expenditure (“diverse recruitment yield”). The results of linear regression analyses weighted by clicks is shown in Figures 2a, 2b and 2c. No significant association was observed between CBSA income and diverse recruitment yield of African American participants (p = 0.99), Latino participants (p = 0.37), or women (p = 0.21) (R^2^ range 0.000–0.13). This analysis was limited to the 14 CBSAs with greater than or equal to 20 recruits. On sensitivity analysis, when we used a cutoff of CBSAs with greater than or equal to 10 or 15 recruits, the conclusions were unchanged (Supplement 5).

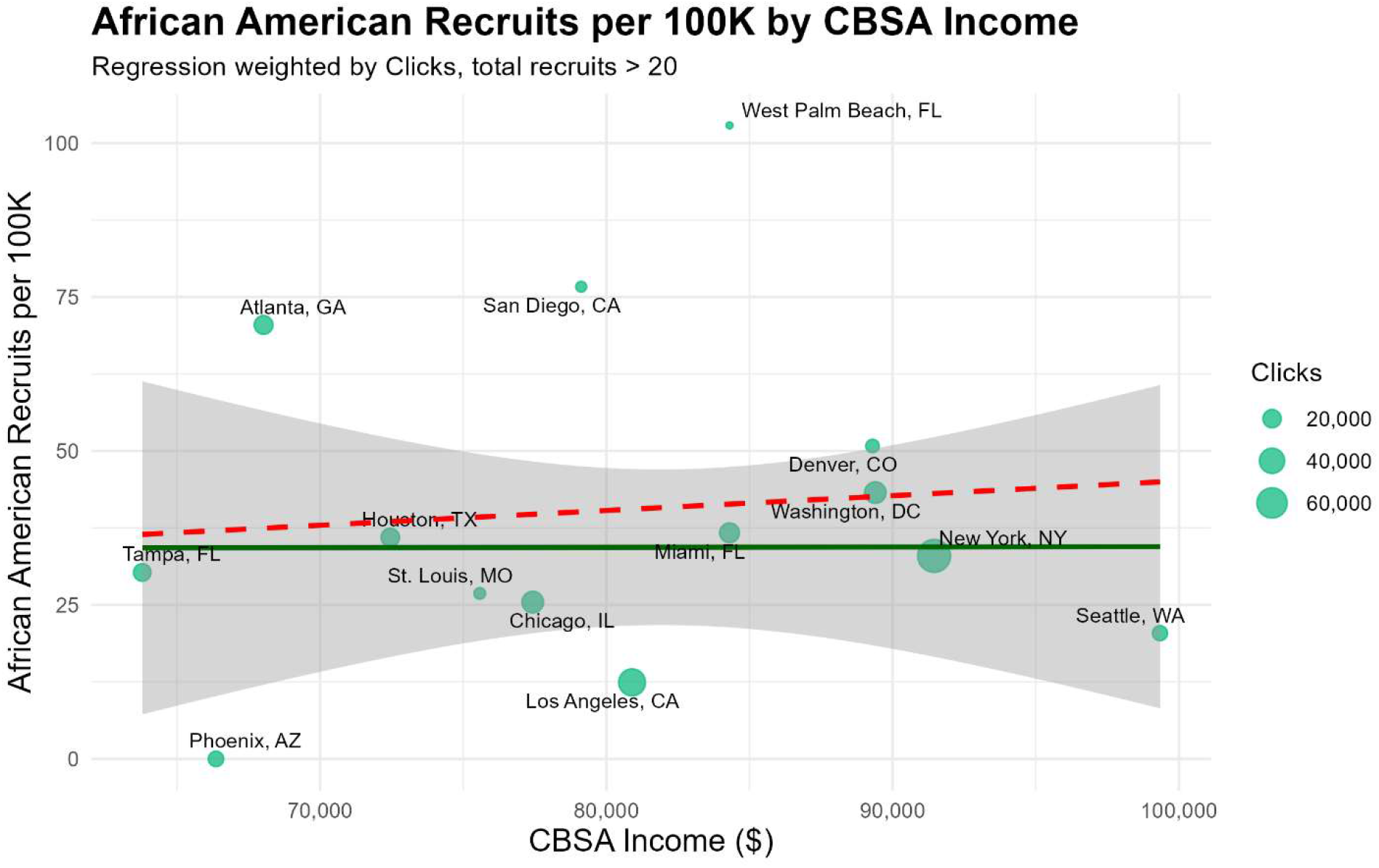
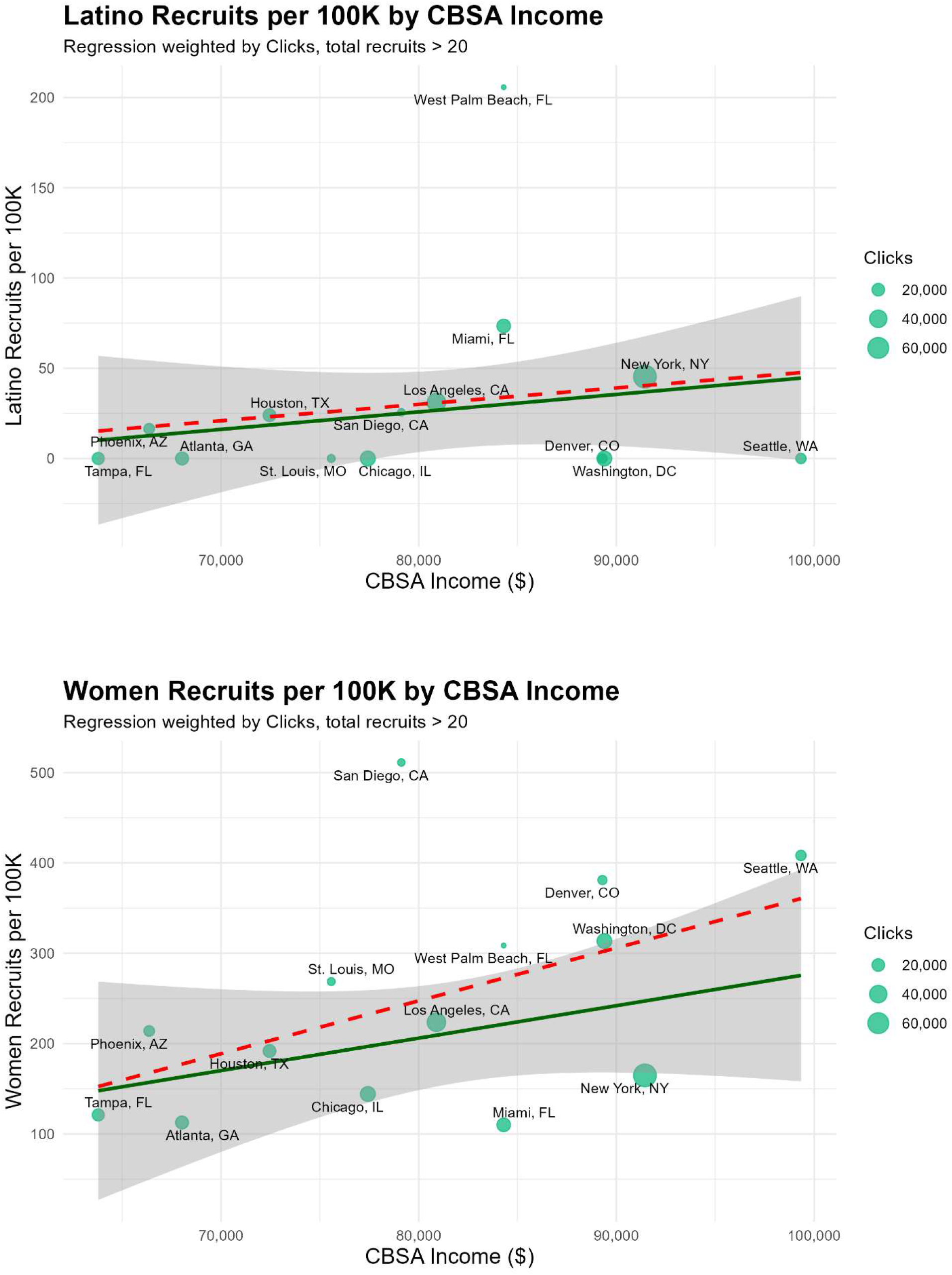
Figure 2a: African American Recruits per 100K by CBSA Income, Figure 2b: Laino Recruits per 100K by CBSA Income and Figure 2c: Women Recruits per 100K by CBSA Income (green line represents weighted regression fit; dotted red line represents unweighted fit).

## Discussion

In this retrospective analysis of national digital advertising data from the Heartbeat Study, we evaluated the feasibility of a digital platform to identify and recruit diverse participants for a randomized clinical trial, LIBREXIA-AF. We also examined how recruitment yield varied among communities stratified by income. We found that our advertising strategy and digital platform were effective in recruiting over 1300 patients from over 260 CBSAs. Despite wide-reaching digital advertising, our recruitment of nonwhite participants remained low (15% of total recruitment). The number of estimated recruits/$100K was 334 for women, 36 for African Americans and 27 for Latinos. There was a significant negative but modest association between CBSA-level income and cost/impression and cost/click suggesting that lower income areas may require slightly more resource input for the same recruitment outcomes. These differences, however, were not seen when we compared expected recruitment yield of diverse groups (African Americans, Latinos, and women) against CBSA income, suggesting that community-level income does not meaningfully predict the ability of digital recruitment strategies to attract diverse participants in cardiovascular clinical trials.

Despite reaching areas of diverse populations via an online strategy, recruitment of nonwhite participants remained low. In 2024, the US Census estimated 58% of the population identified as white, 14% identified as Black, 20% identified as Hispanic/Latino, and 7% identified as Asian.^28^ Even after accounting for a lower rate of AF in non-white populations,^29^ all nonwhite groups were underrepresented in our recruitment, especially Latinos, who only represented 2% of our total survey and possibly limited by English-only recruitment materials. This may have been due in part to our advertising model, which focused materials based on geographic targets, rather than race/ethnicity targets. One trial to promote the participation of African Americans to the National Lung Screening Trial (NSLT) found that after implementing targeted recruitment strategies led to the recruitment of 77.6% (1223 total) of their minority participants. Recruitment strategies included a focus on academic institutions with historically diverse patient populations, community outreach, implementing translations of research materials to Spanish and Asian dialects, and racial/ethnic matching of NLST staff and trial participants.^30^ Potentially, a more targeted approach may have enhanced our recruitment diversity.

Of note, the limited literature that directly reports cost/recruit varies significantly in recruitment costs per enrollment; our cost per recruit of $116.61 appears reasonable; one study of general practice patients selected through EMR review and subsequent research assistant intake reported a cost of $277/recruit with a range of $74-797/recruit across study sites.^31^ Another study of patient recruitment in an aspirin administration clinical trial showed that with a variety of recruitment strategies, email was the most cost effective ($95.71/enrollment) vs in-person ($417.12/enrollment) and letter ($542.26/enrollment).^32^ Further exploration and report on digital strategies especially in the recruitment of underserved participants is certainly needed.

Importantly, among the 691 participants who were eligible for referral to LIBREXIA-AF, only 11 ultimately declined trial participation. This suggested that generating initial engagement among diverse populations may be more important than addressing hesitation to trial enrollment itself. This conclusion reflects the current literature; numerous trials have noted that diverse participants are equally, if not more, likely to participate in trials once asked.^33–35^ Additional factors associated with successful diverse trial recruitment that could be supported by online approaches include publishing materials in a native language (38% of US Hispanics identify as Spanish-dominant vs bilingual or English-dominant^36^), and providing reminders.^37^ Other non-digital approaches include maximizing community and primary care engagement, broadening inclusion/exclusion criteria, and shortening consents and trial duration.^38^

We observed a modest inverse relationship between CBSA-level income and advertising cost per click suggesting that recruitment efforts targeting lower-income communities may require greater advertising expenditures. Lower-income populations are frequently underrepresented in clinical trials, likely due to barriers such as limited access to healthcare, transportation challenges, and greater time constraints.^39,40^ Interestingly, community-level income was not specifically associated with the recruitment yield of underrepresented groups in our analysis. This finding suggests that diverse, lower income individuals may be recruited digitally without significantly higher resource needs than diverse participants from higher income communities. Furthermore, certain cities, such as West Palm Beach, FL and San Diego, CA, demonstrated comparatively higher yield for diverse recruitment at a fixed advertising expenditure. These communities may represent promising opportunities for targeted digital outreach aimed at improving participant diversity in future cardiovascular clinical trials.

Our study had several limitations. As a retrospective analysis, we were constrained to advertising performance data from a single clinical trial recruitment platform which may limit generalizability to other recruitment strategies or populations. Additionally, the digital platform only allowed for advertisement in markets adjacent to sites which had opted-in to external referrals for LIBREXIA-AF recruitment. Secondly, our estimates of “diverse recruitment yield” assumed that the number of diverse participants could be extended proportionally with continued advertising expenditure. In practice, recruitment efficiency may not scale linearly with increased advertisement spend once responsive populations are reached. Thirdly, as we did not capture individual/household income, we relied on CBSA-level income which may be imprecise and may not capture within-community heterogeneity in demographics and digital engagement patterns. Finally, our English-only surveys with multiple web pages may have limited participation among individuals with lower digital literacy as well as non-English language preferences.

## Conclusion

In this national analysis of a digital recruitment platform supporting a large cardiovascular clinical trial, online advertising enabled engagement with participants across a wide range of U.S. communities and socioeconomic settings. Although modest differences in advertising costs were observed across communities of varying income, community-level income was not associated with recruitment yield among women, African American, or Latino participants. Despite broad geographic reach and high overall engagement, racial and ethnic minority representation remained limited, highlighting that digital recruitment alone may be insufficient to meaningfully improve trial diversity. These findings suggest that digital platforms may serve as scalable tools to expand trial reach, but targeted, culturally and linguistically tailored strategies beyond geographic advertising will likely be necessary to substantially improve the enrollment of underrepresented populations in cardiovascular clinical trials.

## Data Availability

Data can be made available upon request.

## References

1. Kelsey MD, Patrick-Lake B, Abdulai R, et al. Inclusion and diversity in clinical trials: Actionable steps to drive lasting change. Contemp Clin Trials. 2022;116:106740. doi:10.1016/j.cct.2022.106740

2. Lan RH, Paranjpe I, Saeed M, Perez MV. Inequities in atrial fibrillation trials: An analysis of participant race, ethnicity, and sex over time. Heart Rhythm. 2025;22(3):602–608. doi:10.1016/j.hrthm.2024.06.052

3. Zhang T, Tsang W, Wijeysundera HC, Ko DT. Reporting and representation of ethnic minorities in cardiovascular trials: a systematic review. Am Heart J. 2013;166(1):52–57. doi:10.1016/j.ahj.2013.03.022

4. Clark LT, Watkins L, Piña IL, et al. Increasing Diversity in Clinical Trials: Overcoming Critical Barriers. Curr Probl Cardiol. 2019;44(5):148–172. doi:10.1016/j.cpcardiol.2018.11.002

5. Melloni C, Berger JS, Wang TY, et al. Representation of Women in Randomized Clinical Trials of Cardiovascular Disease Prevention. Circ Cardiovasc Qual Outcomes. 2010;3(2):135–142. doi:10.1161/CIRCOUTCOMES.110.868307

6. Nunes JC, Rice EN, Stafford RS, Lewis EF, Wang PJ. Underrepresentation of Ethnic and Racial Minorities in Atrial Fibrillation Clinical Trials. Circ Arrhythm Electrophysiol. 2021;14(12). doi:10.1161/CIRCEP.121.010452

7. Tan RKJ, Wu D, Day S, et al. Digital approaches to enhancing community engagement in clinical trials. NPJ Digit Med. 2022;5(1):37. doi:10.1038/s41746-022-00581-1

8. Van Norman GA. Decentralized Clinical Trials: The Future of Medical Product Development?*. JACC Basic Transl Sci. 2021;6(4):384–387. doi:10.1016/j.jacbts.2021.01.011

9. Fisher-Hoch SP, Below JE, North KE, McCormick JB. Challenges and strategies for recruitment of minorities to clinical research and trials. J Clin Transl Sci. 2023;7(1):e154. doi:10.1017/cts.2023.559

10. Banda DR, Libin AV, Wang H, Swain SM. A pilot study of a culturally targeted video intervention to increase participation of African American patients in cancer clinical trials. The Oncologist. 2012;17(5):708–714. doi:10.1634/theoncologist.2011-0454

11. Tang W, Wei C, Cao B, et al. Crowdsourcing to expand HIV testing among men who have sex with men in China: A closed cohort stepped wedge cluster randomized controlled trial. PLoS Med. 2018;15(8):e1002645. doi:10.1371/journal.pmed.1002645

12. Hlatshwako T, Conserve D, Day S, Reynolds Z, Weir S, Tucker JD. Increasing Men’s Engagement in HIV Testing and Treatment Programs Through Crowdsourcing: A Mixed-Methods Analysis in Eswatini. Sex Transm Dis. 2021;48(10):789–797. doi:10.1097/OLQ.0000000000001408

13. Colom A. Using WhatsApp for focus group discussions: ecological validity, inclusion and deliberation. Qual Res. 2022;22(3):452–467. doi:10.1177/1468794120986074

14. Oliffe JL, Kelly MT, Gonzalez Montaner G, Yu Ko WF. Zoom Interviews: Benefits and Concessions. Int J Qual Methods. 2021;20:16094069211053522. doi:10.1177/16094069211053522

15. Kasahara A, Mitchell J, Yang J, Cuomo RE, McMann TJ, Mackey TK. Digital technologies used in clinical trial recruitment and enrollment including application to trial diversity and inclusion: A systematic review. Digit Health. 2024;10:20552076241242390. doi:10.1177/20552076241242390

16. Makri A. Bridging the digital divide in health care. Lancet Digit Health. 2019;1(5):e204–e205. doi:10.1016/S2589-7500(19)30111-6

17. Diversity Action Plans to Improve Enrollment of Participants from Underrepresented Populations in Clinical Studies; Guidance for Industry - Draft Guidance. Regulations.gov. https://www.regulations.gov/document/FDA-2021-D-0789-0111

18. Vargas E, Scherer L, Fiske S. Population Data and Demographics in the United States; “Advancing Antiracism, Diversity, Equity, and Inclusion in STEMM Organizations: Beyond Broadening Participation.”

19. Boxall C, Renz S, Stuart B, et al. Social media recruitment enhances participant diversity in dermatology clinical trial: findings from the SAFA trial. Trials. 2025;26(1):318. doi:10.1186/s13063-025-08994-5

20. Tomiwa T, Wong E, Miller HN, et al. Leveraging digital tools to enhance diversity and inclusion in clinical trial recruitment. Front Public Health. 2024;12:1483367. doi:10.3389/fpubh.2024.1483367

21. Jain SS, Mahaffey KW, Pieper KS, et al. Milvexian vs apixaban for stroke prevention in atrial fibrillation: The LIBREXIA atrial fibrillation trial rationale and design. Am Heart J. 2024;277:145–158. doi:10.1016/j.ahj.2024.08.011

22. A Study of Milvexian Versus Apixaban in Participants With Atrial Fibrillation (LIBREXIA-AF). ClinicalTrials.gov. Accessed March 8, 2026. https://clinicaltrials.gov/study/NCT05757869

23. Jain SS, Mahaffey KW, Pieper KS, et al. Milvexian vs apixaban for stroke prevention in atrial fibrillation: The LIBREXIA atrial fibrillation trial rationale and design. Am Heart J. 2024;277:145–158. doi:10.1016/j.ahj.2024.08.011

24. Delineation Files. United States Census Bureau. doi: https://www.census.gov/geographies/reference-files/time-series/demo/metro-micro/delineation-files.html

25. HUD USPS ZIP Code Crosswalk Files. U.S. Department of Housing and Urban Development. doi:https://www.huduser.gov/apps/public/uspscrosswalk/home

26. US ZIP codes to CBSA. Stanford Medicine Center for Population Health Sciences. https://stanfordphs.redivis.com/datasets/dbhp-0vj8t27f9/tables

27. Personal Income by County. US Bureau of Economic Analysis. https://www.bea.gov/data/income-saving/personal-income-by-county

28. QuickFacts. United States Census Bureau. https://www.census.gov/quickfacts/fact/table/US/PST045224

29. Amponsah MKD, Benjamin EJ, Magnani JW. Atrial Fibrillation and Race - A Contemporary Review. Curr Cardiovasc Risk Rep. 2013;7(5). doi:10.1007/s12170-013-0327-8

30. Duda C, Mahon I, Chen MH, et al. Impact and costs of targeted recruitment of minorities to the National Lung Screening Trial. Clin Trials. 2011;8(2):214–223. doi:10.1177/1740774510396742

31. Tew M, Catchpool M, Furler J, et al. Site-specific factors associated with clinical trial recruitment efficiency in general practice settings: a comparative descriptive analysis. Trials. 2023;24(1):164. doi:10.1186/s13063-023-07177-4

32. Raby K, Blinson K, Herrington DM, et al. Abstract 14173: Emailing is an Economical Way to Recruit Participants. Circulation. 2020;142(Suppl_3). doi:10.1161/circ.142.suppl_3.14173

33. Thetford K, Gillespie TW, Kim YI, Hansen B, Scarinci IC. Willingness of Latinx and African Americans to Participate in Nontherapeutic Trials: It Depends on Who Runs the Research. Ethn Dis. 2021;31(2):263–272. doi:10.18865/ed.31.2.263

34. Adeyemi OF, Evans AT, Bahk M. HIV-infected adults from minority ethnic groups are willing to participate in research if asked. AIDS Patient Care STDs. 2009;23(10):859–865. doi:10.1089/apc.2009.0008

35. Langford AT, Resnicow K, Dimond EP, et al. Racial/ethnic differences in clinical trial enrollment, refusal rates, ineligibility, and reasons for decline among patients at sites in the National Cancer Institute’s Community Cancer Centers Program. Cancer. 2014;120(6):877–884. doi:10.1002/cncr.28483

36. Language Use Among Latinos. Pew Research Center. https://www.pewresearch.org/race-and-ethnicity/2012/04/04/iv-language-use-among-latinos/

37. Treweek S, Pitkethly M, Cook J, et al. Strategies to improve recruitment to randomised trials. Cochrane Methodology Review Group, ed. Cochrane Database Syst Rev. 2018;2018(2). doi:10.1002/14651858.MR000013.pub6

38. Barriers to Representation of Underrepresented and Excluded Populations in Clinical Research; from Improving Representation in Clinical Trials and Research: Building Research Equity for Women and Underrepresented Groups.

39. Donzo MW, Nguyen G, Nemeth JK, et al. Effects of socioeconomic status on enrollment in clinical trials for cancer: A systematic review. Cancer Med. 2024;13(1):e6905. doi:10.1002/cam4.6905

40. Unger JM, Gralow JR, Albain KS, Ramsey SD, Hershman DL. Patient Income Level and Cancer Clinical Trial Participation: A Prospective Survey Study. JAMA Oncol. 2016;2(1):137–139. doi:10.1001/jamaoncol.2015.3924

